# The Burden of Hospital Morbidity and Mortality in Ghana: A Four-Year Retrospective Analysis of National Data

**DOI:** 10.64898/2026.01.13.26344071

**Authors:** Alfred Kwesi Manyeh, Christopher Tetteh Odopey, Angela Nana Esi Ackon, Vincent Uwumboriyhie Gmayinaam, Patrick Dzissem, Evelyn Acquah, Mustapha Immurana

## Abstract

**Background:** Understanding hospital admission and mortality trends is essential for enhancing health system performance in low- and middle-income countries. In Ghana, systemic challenges such as unequal access to care, workforce shortages, and changing disease patterns – exacerbated by the COVID-19 pandemic – underscore the need for comprehensive health data analysis. This study examines national hospital morbidity and mortality trends from 2019 to 2023, identifying demographic and clinical factors that predict mortality to inform policy and health system reforms.

**Methods:** A retrospective analysis was performed using data from Ghana’s District Health Information Management System-2 (DHIMS-2), covering 2,357,535 hospital admissions across 15 regions from 2019 to 2023. Descriptive statistics and multivariate logistic regression were employed to investigate patterns in morbidity, mortality, and associated risk factors, based on patient demographics, insurance status, disease classification (ICD-11), and length of hospital stay.

**Results:** Hospital mortality throughout the study period was 2.6%. Infectious and parasitic diseases were the primary causes of admission (30.1%), while diseases of the nervous system exhibited the highest case fatality rate (13.8%). Mortality was significantly greater from 2020 to 2023 compared with 2019, with the highest odds observed in 2023 (AOR = 1.50; 95% CI: 1.46–1.54). Advanced age (≥65 years), male sex, lack of insurance, short hospital stays (<1 day), and comorbid conditions were all linked to higher mortality. Admissions due to neoplasms, circulatory, respiratory, and neurological conditions demonstrated notably high adjusted odds of mortality. Conversely, admissions covered by health insurance, female patients, and those with stays of 3–7 days had a reduced mortality risk.

**Conclusion:** This national review highlights ongoing gaps in the health system that lead to hospital mortality, especially for complex conditions and vulnerable groups. While the National Health Insurance Scheme looks protective, disparities still exist across age, sex, region, and disease types. Urgent investments in diagnostic and critical care, along with expanding insurance coverage and regional resource distribution, are needed to reduce preventable hospital deaths in Ghana.

## Background

One well-known measure of the effectiveness of the healthcare system and the standard of treatment provided is hospital mortality. The clinical burden of illness, the quality of care provided, and the broader socioeconomic factors influencing health outcomes are all reflected in it. Hospital mortality is still too high in many low- and middle-income countries (LMICs), primarily due to non-communicable and communicable diseases, health system limitations, and delayed health-seeking behaviours [1], [2].

In LMICs, especially in sub-Saharan Africa, infectious diseases like HIV/AIDS, malaria, pneumonia, and tuberculosis remain among the top causes of hospital fatalities [2], [3]. Non-communicable diseases (NCDs), such as cancer, diabetes, cardiovascular disease, and chronic respiratory conditions, are, nevertheless, contributing more and more to in-hospital mortality as a result of the ongoing epidemiological transition. These diseases frequently require specialised and prolonged care, which many medical facilities are poorly equipped to provide [3], [4].

Numerous individual-level factors are consistently associated with an increased risk of hospital mortality. These include older age, due to physiological decline and multimorbidity, as well as male sex, often linked with poorer health-seeking behaviour and a higher prevalence of high-risk conditions; and low socioeconomic status, which affects access to healthcare and the ability to manage chronic illness effectively [5], [6], [7]. Delayed presentation to health facilities, sometimes due to cultural practices or geographic inaccessibility, also exacerbates patient outcomes, particularly in emergency or time-sensitive conditions [8].

It has been demonstrated that not having health insurance greatly lowers the likelihood of seeking care as soon as possible, which results in worse admission prognoses. Research from Ghana and other African nations indicates that individuals with insurance experience better health outcomes and are more likely to utilise preventive and curative care [9].

Health system factors, such as shortages of healthcare workers, limited diagnostic capacity, overburdened referral systems, and a lack of critical care infrastructure, particularly in rural or peripheral health facilities, significantly contribute to hospital mortality [10].

Ultimately, hospital mortality is not only a function of disease severity but also a reflection of systemic inefficiencies. Addressing it requires integrated approaches: improving community-level disease surveillance, strengthening health worker training, ensuring equitable insurance coverage, and expanding diagnostic and emergency care infrastructure. Generating evidence on the distribution and determinants of hospital mortality remains essential for guiding national health policies and achieving universal health coverage [11].

### Main Objective

To analyse the trends and spatial distribution of morbidity and causes of death in Ghana, informing targeted health interventions, evidence-based policy development, and effective resource allocation for improved healthcare outcomes.

### Specific Objectives

1. To analyse the trend of morbidity in Ghana over a specified period. (2019-2023)
2. To examine the spatial distribution of causes of death in Ghana.
3. To identify areas with high morbidity and mortality rates.
4. To determine the leading causes of death in Ghana.

## Methods

### Study design

A retrospective analysis of nationally representative data from 2,357,535 hospital admissions that occurred during the five years from 2019 to 2023, across 15 of the 16 regions of Ghana, was conducted (excluding the North East Region, which had incomplete data for several years).

### Data source

Data was extracted from the Ghana Health Service’s District Health Information Management System-2 (DHIMS-2) database. Except for the leading teaching hospitals, which report specific indicators, the DHIMS-2 has data on all patients admitted to public and private health facilities in Ghana. It is currently the country’s most comprehensive national database of routine health data [12].

#### Study site

Data for this study covered fifteen (15) administrative regions of Ghana, as shown in Figure 1. Ghana is a country in West Africa, covering an area of 238,537 km² (92,099 square miles) with a population of approximately 31 million. It is the second-most populous country in West Africa, after Nigeria; Accra is its capital and largest city [13]. The population of Ghana, according to the 2021 national census, is 30,832,019, with 56.7% of the population residing in urban areas [14]. Health services are delivered in public and private health facilities, which are supervised by the Ghana Health Service. The provision of health services in Ghana ranges from the highest of tertiary hospitals to the lowest of Community-Based Health Planning and Services. Currently, there are approximately 6,000 health facilities providing health services in Ghana, with around 75% being government-owned. The two largest cities, Accra (the national capital) and Kumasi (the regional capital of the Ashanti Region), have the highest concentration of health facilities and doctors in the country. It is estimated that approximately 66.2% of individuals who report being ill or injured consult a healthcare practitioner in Ghana. Overall, 67.6 % of the population is registered or covered by the health insurance scheme [14].

**Figure 1:**
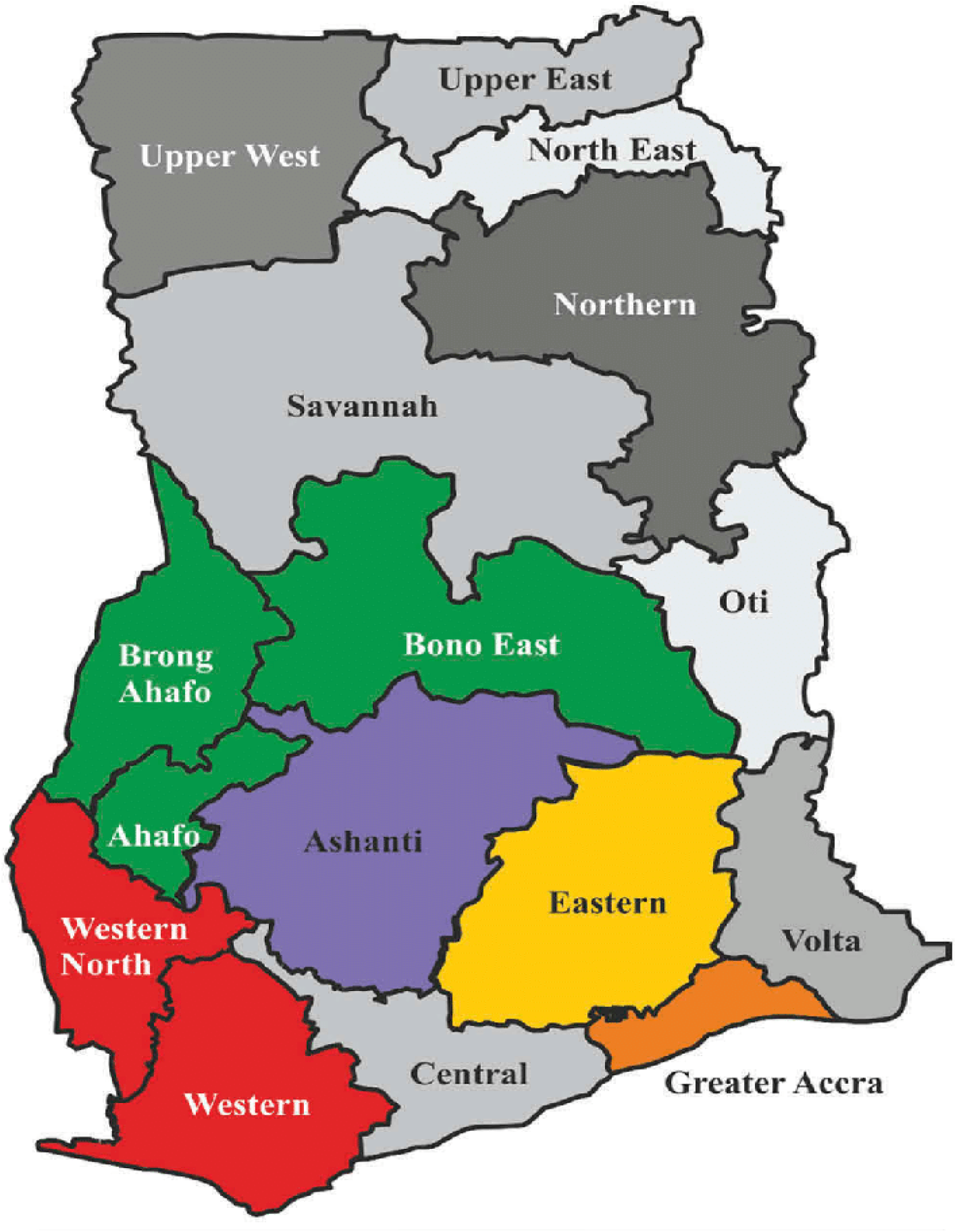
The Map of Ghana Showing the Sixteen (16) Regions. [15]

#### Data extraction and variable measurement

Data on hospital admissions covering the study period in the DHIMS-2 database were extracted and exported into Microsoft Excel for cleaning. The explanatory variables include year of admission, age, occupation, marital status, level of education, type of health facility, health facility status, region, and health insurance status. The outcome variable is the admission outcome, which was dichotomous (coded as 1 if there was mortality and 0 if not). The overall mortality rate in this study was calculated as a percentage by dividing the total number of mortalities by the total number of hospital admissions over the study period (2019-2023).

#### Data Analysis

**Morbidity:** Refers to the number of hospital admission cases broadly classified using the ICD-11 coding tool.

**Mortality:** Refers to the number of deaths resulting from the admitted cases.

**Case fatality:** Refers to the proportion of morbidities, classified using ICD-11 broad classification, which resulted in mortality with respect to that specific category.

The data was exported into Microsoft Excel 2021 for cleaning by removing blank cells. The data were then exported into Stata/MP version 17 for statistical analysis. Data was analysed descriptively by person, place and time using frequencies and proportions. To measure the odds of mortality, logistic regression analysis was done at two levels. The first level was to establish the crude level of association between mortality and the independent variables for the study. For this study, all independent variables were added to the model to develop the crude relationship with mortality. All independent variables with p < 0.05 in the bivariate analysis were included in the multivariate logistic regression further to examine the association between mortality and each independent variable, while controlling for the effect of other variables by calculating adjusted odds ratios (AOR). The level of significance used was 5% (0.05), two-tailed, with a 95% confidence interval (CI). Results are presented using tables, charts and graphs.

## Results

A total of 2,357,535 hospital admissions occurred nationally (excluding the North East Region, which had incomplete data for several years) during the five years from 2019 to 2023. Regarding years, 2019 accounted for 24.0% of the total admissions for the study period, 2020 for 21.4%, 2022 for 21.1%, 2021 for 17.4%, and 2023 for the lowest at 16.1% of the hospital admissions. Based on regions, the Eastern region recorded the highest proportion (19.2%) of hospital admissions during the study period. Greater Accra and Ashanti regions each recorded 16.5%. The rest of the areas each recorded less than 10% of hospital admissions. Based on the cause of morbidity using broad ICD-11 classifications, infectious or parasitic diseases accounted for the highest (30.1%) cause of morbidity resulting in hospital admissions. Pregnancy, childbirth or the puerperium accounted for 14.7% of hospital admissions, diseases of the respiratory system resulted in 7.7% of hospital admissions, diseases of the digestive system had 7.2%, certain conditions originating in the perinatal period had 6.2%, diseases of the genitourinary system had 5.9%, diseases of the circulatory system had 5.6%, diseases of the blood or blood-forming organs recorded 5.0% and the rest of the ICD-11 classifications each recorded less than 4% of hospital admissions. Regarding the outcome of hospital admissions, the majority (94.6%) were discharged, 2.6% died, 1.7% were referred, 0.5% absconded, and 0.6% of the admission outcomes were unspecified. Based on the presence of another diagnosis in addition to the primary diagnosis, more than half (73.5%) of hospital admissions did not have another diagnosis, while the rest (26.5%) had one. The mean number of days of hospitalisation was 3.5±8.1, with 34.9% recording 3-7 days of hospitalisation, 27.9% recording 2 days, 22.3% recording 1 day, and 7.4% each recording less than 1 day and more than 1 week. Based on insurance status, more than half (81.7%) of respondents reported being insured, while the rest were not insured. Females comprised the majority (61.1%) of the respondents, while the remainder were males. The mean age of respondents was 28±22.8 with 6.4% of the respondents less than 1 year old, 15.3% were aged 1-4 years, 11.3% were aged 5-14 years, (15.1%) were 15-24 years old, 18.4% were aged 25-34 years, 11.6% were aged 35-44 years, 7.3% were 45-54 years old, 6.0% were aged 55-64 years and the rest were 65 years and above. Regarding education, 45.7% had no formal education, while the rest had some form of education. Based on occupation, 38.5% were either students or children, 32.5% were employed, 6.8% were unemployed, 2.2% were retired, and 20.0% were not specified (see Table 1).

**Table 1:** Distribution of hospital admissions from 2019 to 2023.

| Variable | Frequency (n = 2,357,535) | Percentage (%) |
| --- | --- | --- |
| **Year** |  |  |
| 2019 | 565,451 | 24.0 |
| 2020 | 504,954 | 21.4 |
| 2021 | 410,340 | 17.4 |
| 2022 | 497,045 | 21.1 |
| 2023 | 379,745 | 16.1 |
| **Region** |  |  |
| Ahafo | 55,768 | 2.4 |
| Ashanti | 387,868 | 16.5 |
| Bono | 44,035 | 1.9 |
| Bono East | 52,710 | 2.2 |
| Central | 145,114 | 6.2 |
| Eastern | 451,593 | 19.2 |
| Greater Accra | 388,210 | 16.5 |
| Northern | 104,039 | 4.4 |
| Oti | 39,325 | 1.7 |
| Savannah | 29,960 | 1.3 |
| Upper East | 22,604 | 1.0 |
| Upper West | 131,314 | 5.6 |
| Volta | 196,621 | 8.3 |
| Western | 232,642 | 9.9 |
| Western North | 75,732 | 3.2 |
| **Outcome of admission** |  |  |
| Discharged | 2,230,864 | 94.6 |
| Referred | 41,145 | 1.7 |
| Died | 61,192 | 2.6 |
| Absconded | 10,935 | 0.5 |
| Unspecified | 13,399 | 0.6 |
| **Cause of morbidity (based on ICD-11 classifications)** |  |  |
| Certain infectious or parasitic diseases | 709,862 | 30.1 |
| Neoplasms | 31,464 | 1.3 |
| Diseases of the blood or blood-forming organs | 118,448 | 5.0 |
| Diseases of the immune system | 184 | 0.0 |
| Endocrine, nutritional or metabolic diseases | 86,249 | 3.7 |
| Mental, behavioural or neurodevelopmental disorders | 16,375 | 0.7 |
| Sleep-wake disorders | 534 | 0.0 |
| Diseases of the nervous system | 56,627 | 2.4 |
| Diseases of the visual system | 6,132 | 0.3 |
| Diseases of the ear or mastoid process | 7,352 | 0.3 |
| Diseases of the circulatory system | 132,590 | 5.6 |
| Diseases of the respiratory system | 182,472 | 7.7 |
| Diseases of the digestive system | 168,932 | 7.2 |
| Diseases of the skin | 5,527 | 0.2 |
| Diseases of the musculoskeletal system/connective tissue | 15,445 | 0.7 |
| Diseases of the genitourinary system | 139,888 | 5.9 |
| Pregnancy, childbirth or the puerperium | 345,977 | 14.7 |
| Certain conditions originating in the perinatal period | 146,598 | 6.2 |
| Developmental anomalies | 5,423 | 0.2 |
| Symptoms, signs or clinical findings,<br>not elsewhere classified | 74,766 | 3.2 |
| Injury, poisoning or certain other<br>consequences of external causes | 80,544 | 3.4 |
| External causes of morbidity or<br>mortality | 25,846 | 1.1 |
| Factors influencing health status or<br>contact with health services | 298 | 0.0 |
| Codes for special purposes | 2 | 0.0 |
| **Other diagnosis** |  |  |
| No | 1,732,418 | 73.5 |
| Yes | 625,117 | 26.5 |
| **Mean number of days on admission**<br>**(SD)** | 3.5(8.1) |  |
| **Number of days on admission** |  |  |
| <1 day | 172,549 | 7.4 |
| 1 day | 522,539 | 22.3 |
| 2 days | 652,649 | 27.9 |
| 3-7 days | 816,894 | 34.9 |
| >1 week | 173,643 | 7.4 |
| **Insurance status** |  |  |
| No | 428,418 | 18.3 |
| Yes | 1,910,638 | 81.7 |
| **Mean age (in years) (SD)** | 27.9(22.8) |  |
| **Age** |  |  |
| <1 year | 151,049 | 6.4 |
| 1-4 years | 358,929 | 15.3 |
| 5-14 years | 265,620 | 11.3 |
| 15-24 years | 354,199 | 15.1 |
| 25-34 years | 432,075 | 18.4 |
| 35-44 years | 273,309 | 11.6 |
| 45-54 years | 171,477 | 7.3 |
| 55-64 years | 140,381 | 6.0 |
| 65 and above | 204,311 | 8.7 |
| **Gender** |  |  |
| Male | 916,703 | 38.9 |
| Female | 1,440,761 | 61.1 |
| **Highest level of education** |  |  |
| None | 1,076,596 | 45.7 |
| Primary | 299,854 | 12.7 |
| JHS/Middle | 512,862 | 21.8 |
| SHS/Vocational/Technical | 307,712 | 13.1 |
| Tertiary | 159,451 | 6.8 |
| **Occupation** |  |  |
| Employed | 766,807 | 32.5 |
| Unemployed | 160,683 | 6.8 |
| Student/Child | 907,050 | 38.5 |
| Retired | 52,008 | 2.2 |
| Not specified | 470,975 | 20.0 |

Table 2 presents the frequency distribution of mortalities among hospital admissions during the study period. Out of 2,357,535 hospital admissions, a total of 61,192, representing 2.6% resulted in mortality. The highest proportion of mortality occurred in 2022, at 22.0%, followed by 2020, at 21.6%. The rest of the years recorded less than 20% and the least mortality for the period occurred in 2019, with 17.2%. Moreover, the Greater Accra region recorded the highest proportion of mortality with 27.7%. The Eastern region recorded 22.9%, the Volta region recorded 10.2%, the Ashanti region recorded 9.8%, the Western region recorded 8.2% and the Central region recorded 5.3%. The rest of the areas each recorded less than 5% of mortalities. Based on broad ICD-11 classifications, 18.6% of the mortalities were due to certain infectious or parasitic diseases, 12.7% were due to diseases of the nervous system, diseases of the circulatory system contributed 11.4%, diseases of the respiratory system contributed 11.2%, certain conditions originating in the perinatal period recorded 9.2%, diseases of the digestive system recorded 6.6%, endocrine, nutritional or metabolic diseases recorded 6.1% and symptoms, signs or clinical findings, not elsewhere classified recorded 5.5%. The rest of the ICD-11-based classifications each contributed less than 5% to the mortalities resulting from hospital admissions.

**Table 2:** Distribution of mortalities among hospital admissions from 2019 to 2023.

| Variable | Frequency (n = 61,192) | Percentage (%) |
| --- | --- | --- |
| **Year** |  |  |
| 2019 | 10,553 | 17.2 |
| 2020 | 13,234 | 21.6 |
| 2021 | 11,962 | 19.5 |
| 2022 | 13,448 | 22.0 |
| 2023 | 11,995 | 19.6 |
| **Region** |  |  |
| Ahafo | 663 | 1.1 |
| Ashanti | 6,010 | 9.8 |
| Bono | 1,163 | 1.9 |
| Bono East | 936 | 1.5 |
| Central | 3,272 | 5.3 |
| Eastern | 14,018 | 22.9 |
| Greater Accra | 16,968 | 27.7 |
| Northern | 2,160 | 3.5 |
| Oti | 622 | 1.0 |
| Savannah | 196 | 0.3 |
| Upper East | 362 | 0.6 |
| Upper West | 2,592 | 4.2 |
| Volta | 6,224 | 10.2 |
| Western | 5,029 | 8.2 |
| Western North | 977 | 1.6 |
| **Cause of mortality (based on ICD-11 classifications)** |  |  |
| Certain infectious or parasitic diseases | 11,370 | 18.6 |
| Neoplasms | 1,494 | 2.4 |
| Diseases of the blood or blood-forming organs | 2,897 | 4.7 |
| Diseases of the immune system | 11 | 0.0 |
| Endocrine, nutritional or metabolic diseases | 3,709 | 6.1 |
| Mental, behavioural or neurodevelopmental disorders | 686 | 1.1 |
| Sleep-wake disorders | 12 | 0.0 |
| Diseases of the nervous system | 7,788 | 12.7 |
| Diseases of the visual system | 54 | 0.1 |
| Diseases of the ear or mastoid process | 45 | 0.1 |
| Diseases of the circulatory system | 6,995 | 11.4 |
| Diseases of the respiratory system | 6,850 | 11.2 |
| Diseases of the digestive system | 4,040 | 6.6 |
| Diseases of the skin | 117 | 0.2 |
| Diseases of the musculoskeletal system/connective tissue | 176 | 0.3 |
| Diseases of the genitourinary system | 2,505 | 4.1 |
| Pregnancy, childbirth or the puerperium | 804 | 1.3 |
| Certain conditions originating in the perinatal period | 5,659 | 9.2 |
| Developmental anomalies | 250 | 0.4 |
| Symptoms, signs or clinical findings, not elsewhere classified | 3,342 | 5.5 |
| Injury, poisoning or certain other consequences of external causes | 2,019 | 3.3 |
| External causes of morbidity or mortality | 363 | 0.6 |
| Factors influencing health status or contact with health services | 6 | 0.0 |
| Codes for special purposes | 0 | 0.0 |
| **Other diagnosis** |  |  |
| No | 40,408 | 66.0 |
| Yes | 20,784 | 34.0 |
| **Mean number of days on admission (SD)** | 4.6(10.9) |  |
| **Number of days on admission** |  |  |
| <1 day | 11,172 | 18.4 |
| 1 day | 13,937 | 23.0 |
| 2 days | 8,784 | 14.5 |
| 3-7 days | 17,045 | 28.1 |
| >1 week | 9,641 | 15.9 |
| **Insurance status** |  |  |
| No | 18,770 | 31.0 |
| Yes | 41,763 | 69.0 |
| **Mean age (in years) (SD)** | 45.7(27.0) |  |
| **Age** |  |  |
| <1 year | 5,829 | 9.5 |
| 1-4 years | 3,121 | 5.1 |
| 5-14 years | 2,472 | 4.0 |
| 15-24 years | 2,710 | 4.4 |
| 25-34 years | 4,718 | 7.7 |
| 35-44 years | 7,391 | 12.1 |
| 45-54 years | 8,835 | 14.5 |
| 55-64 years | 8,861 | 14.5 |
| 65 and above | 17,134 | 28.1 |
| **Gender** |  |  |
| Male | 32,190 | 52.6 |
| Female | 29,000 | 47.4 |
| **Highest level of education** |  |  |
| None | 26,765 | 43.8 |
| Primary | 6,164 | 10.1 |
| JHS/Middle | 16,113 | 26.4 |
| SHS/Vocational/Technical | 8,351 | 13.7 |
| Tertiary | 3,742 | 6.1 |
| **Occupation** |  |  |
| Employed | 21,651 | 35.4 |
| Unemployed | 5,436 | 8.9 |
| Student/Child | 12,210 | 20.0 |
| Retired | 5,771 | 9.4 |
| Not specified | 16,112 | 26.3 |

Furthermore, more than half (66.0%) of the total mortalities did not have another diagnosis aside from the primary diagnosis that resulted in admission. The mean number of days on admission for mortalities was 4.6±10.9, with 28.1% of the mortalities spending 3-7 days on admission, 23.0% recording 1 day, and the remaining length of hospital stay each recording less than 20% of mortalities, respectively. Regarding insurance status, 69.0% of the mortalities had insurance, while the rest had no insurance. Males constituted 52.6% of the mortalities, and 47.4% were females. The mean age of mortalities was 45.7±27.0 with the highest proportion (28.1%) of mortality occurring among those aged 65 years and above and the least proportion (4.0%) occurring among those aged 5-14 years. Based on education, 43.8% of the mortalities had no education and the rest had some form of education. Similarly, 35.4% of the mortalities were employed, 26.3% did not have their occupation specified, 20.0% were either a student or child, 9.4% were retired and 8.9% were unemployed, see Table 2.

Table 3 presents the case-fatality rate based on broad ICD classifications. Diseases of the nervous system had the highest case-fatality rate, with 13.8% of hospital admissions from this category resulting in mortality. Diseases of the immune system recorded 6.0%, diseases of the circulatory system recorded 5.3%, neoplasms had 4.8%, developmental anomalies had 4.6%, symptoms, signs or clinical findings, not elsewhere classified recorded 4.5%, endocrine, nutritional or metabolic diseases recorded 4.3% and mental, behavioural or neurodevelopmental disorders recorded 4.2% mortalities out of the total admissions due to their respective categories. The rest of the ICD-11 classifications recorded a case fatality rate of less than 4%, including diseases of the visual system, diseases of the ear or mastoid process, pregnancy, childbirth, or the puerperium, and codes for special purposes, each with a mortality rate of less than 1% from hospital admissions related to their respective categories.

**Figure 2:**
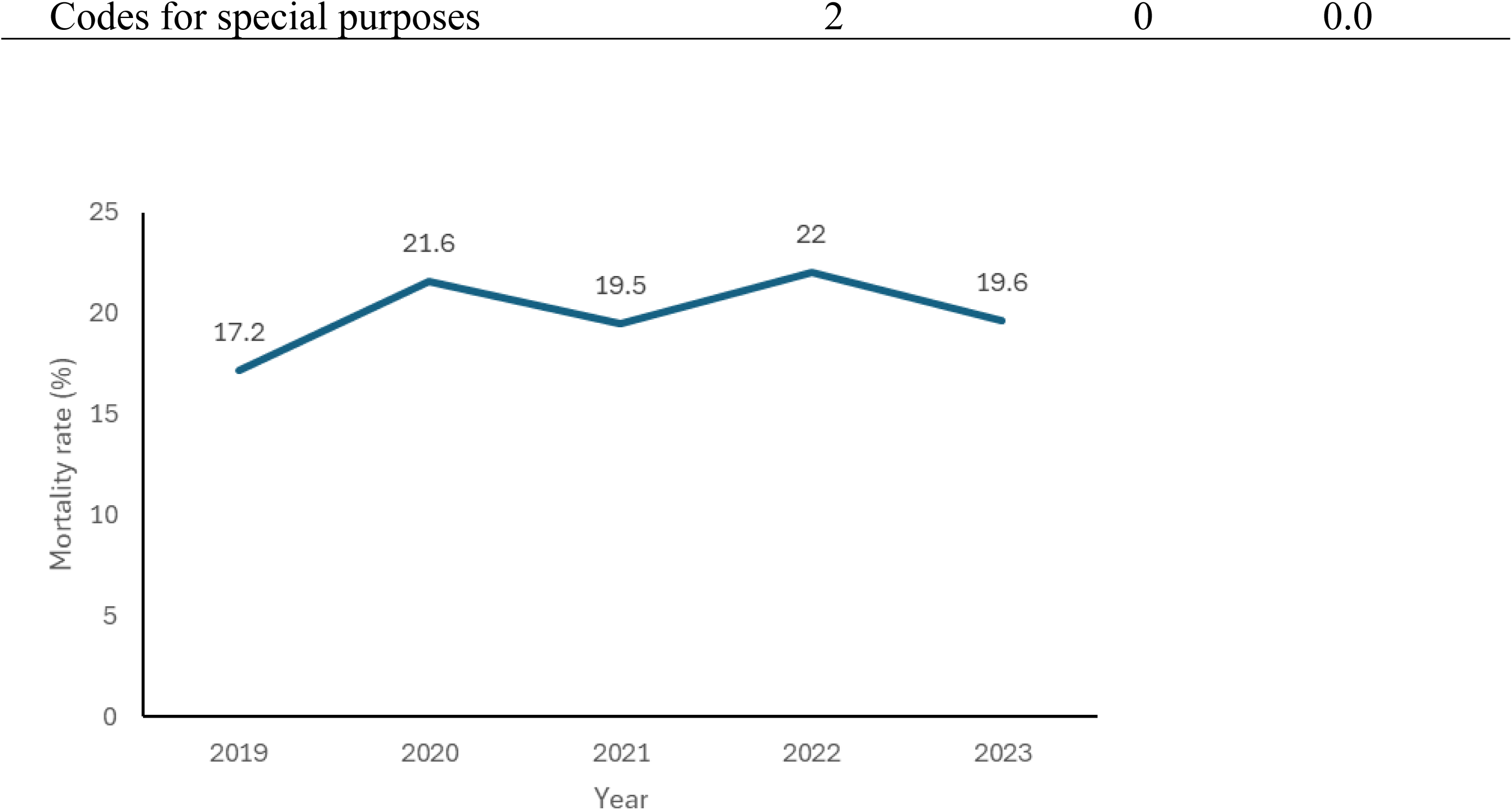
Trends in mortality rate resulting from hospital admissions in Ghana, 2019 to 2023

**Figure 3:**
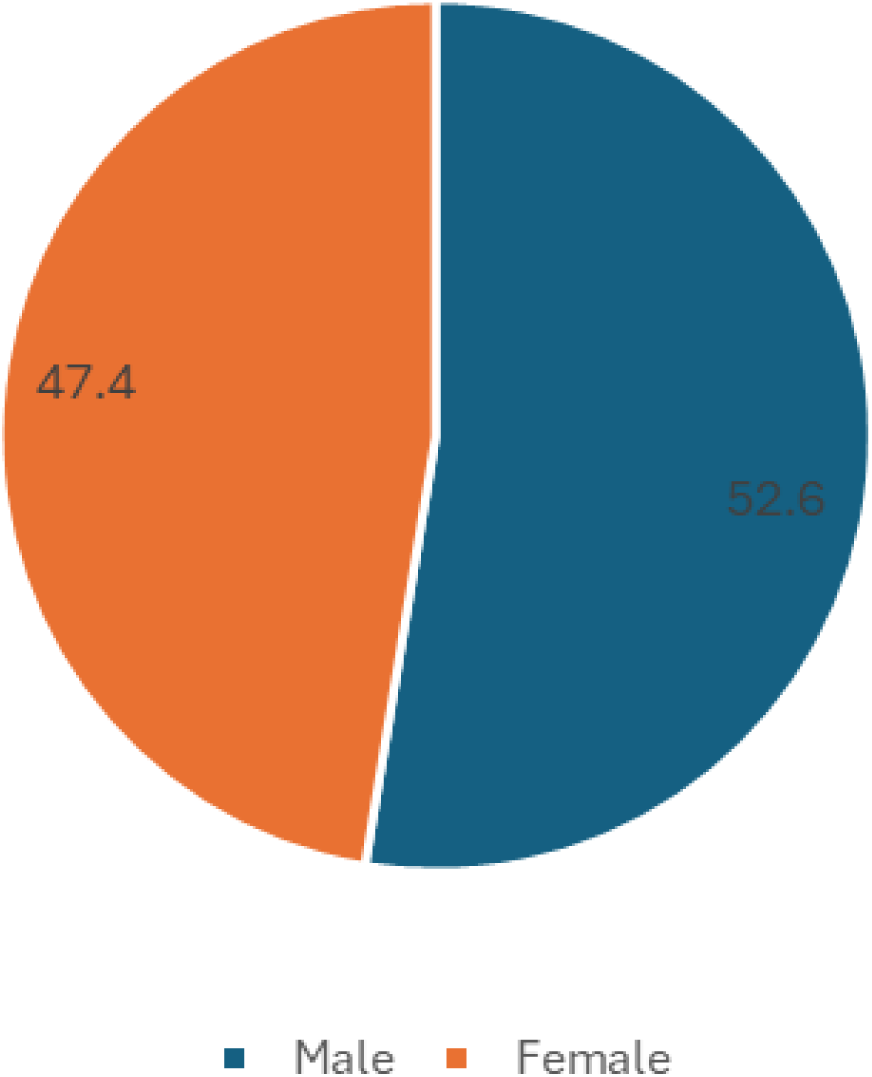
Percentage distribution of mortality by gender

**Figure 4:**
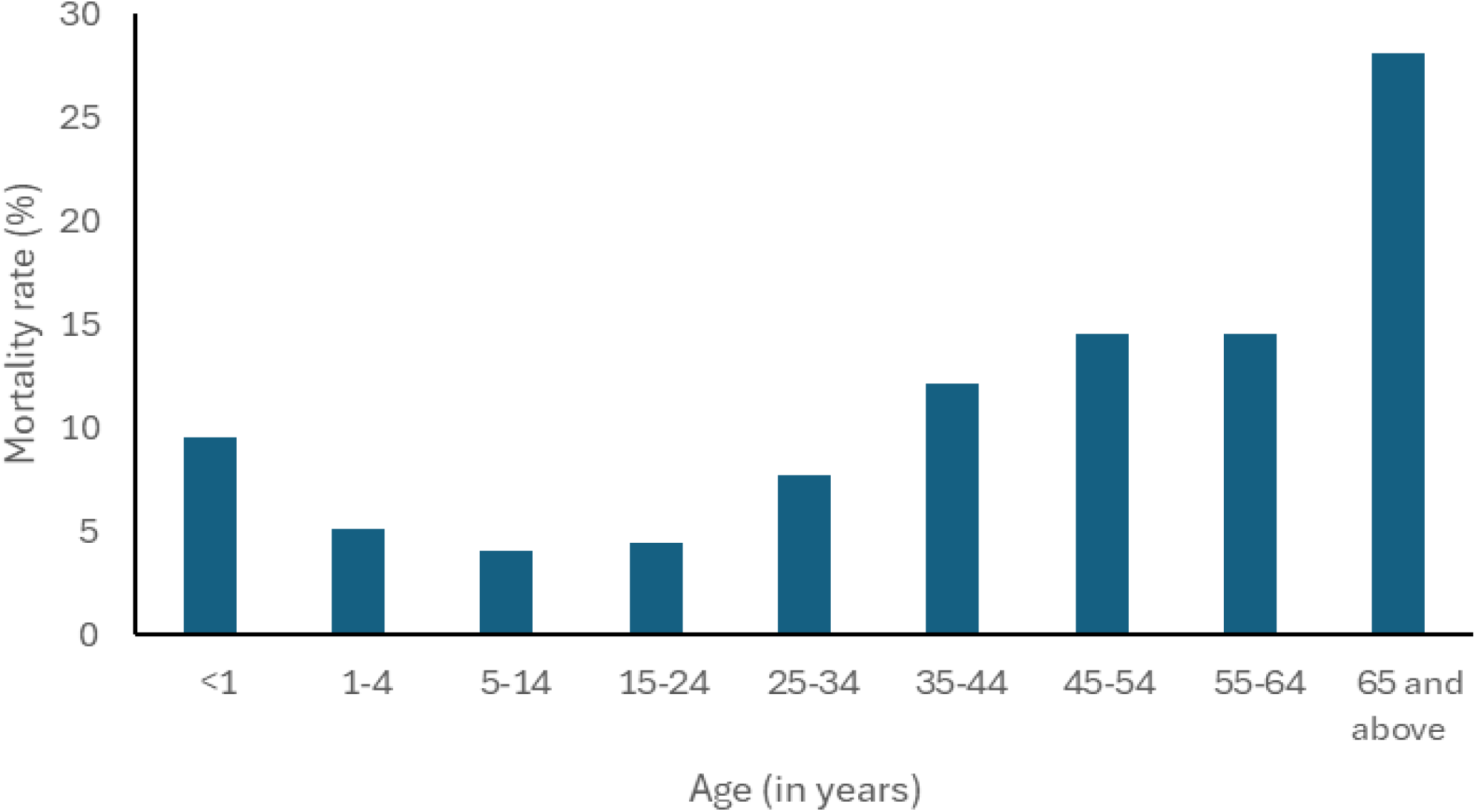
Distribution of mortality by age

**Figure 5:**
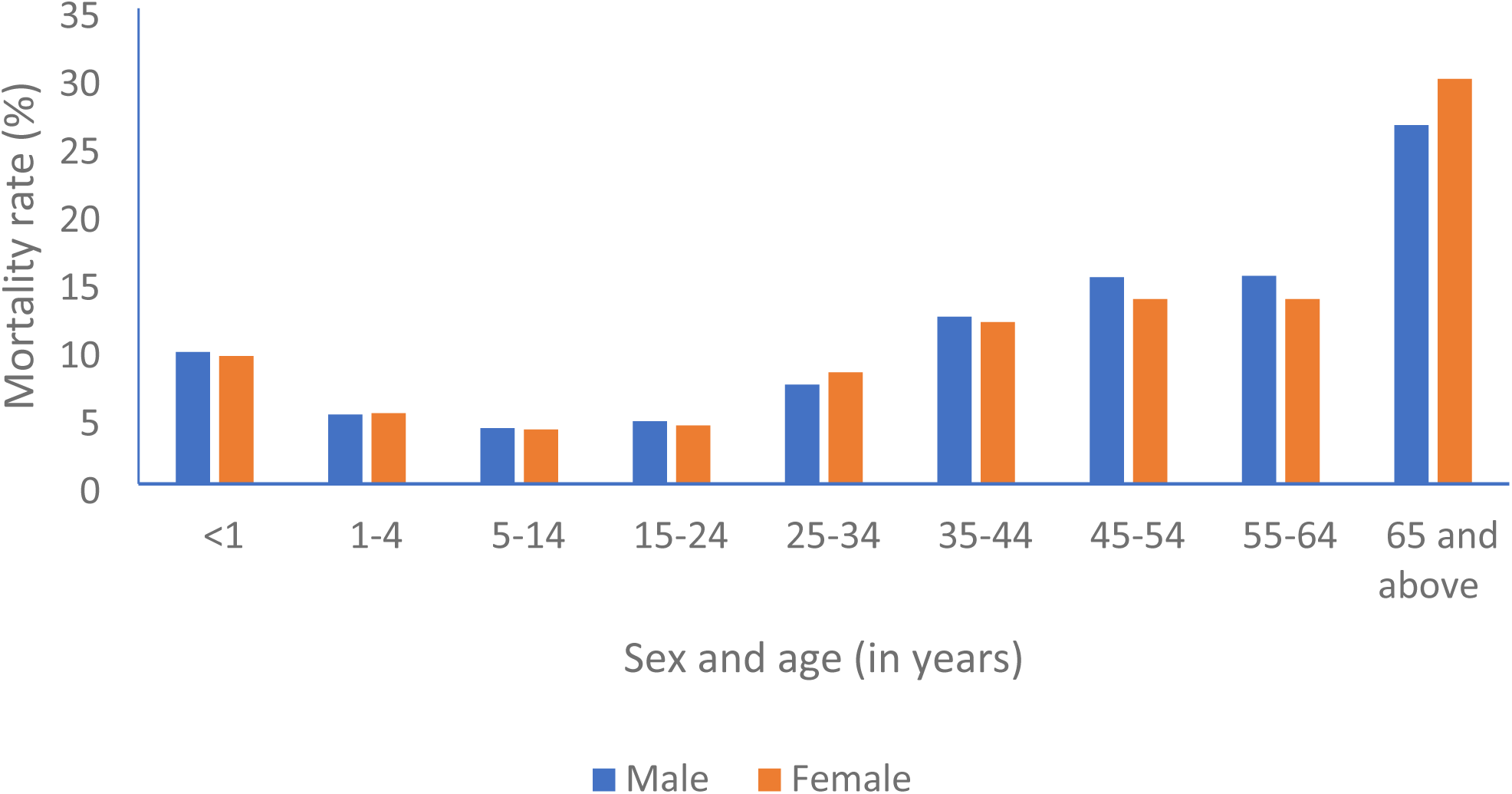
Distribution of mortalities by sex and age Multivariable logistic regression was used to identify factors associated with mortality among hospital admissions. In the adjusted analysis, admissions in 2020, 2021, 2022, and 2023 had significantly higher odds of mortality than hospital admissions in 2019. Hospital admissions in 2020 were 1.29 times more likely to result in mortality (AOR=1.29; 95% CI: 1.26-1.33; p<0.001), admissions in 2021 were 1.27 times more likely (AOR=1.27; 95% CI: 1.23-1.30; p<0.001), admissions in 2022 were 1.30 times more likely (AOR=1.30; 95% CI: 1.26-1.33; p<0.001) and admissions in 2023 were 1.5 times more likely (AOR=1.50; 95% CI: 1.46-1.54; p<0.001) to result in mortality as compared to hospital admissions in 2019.

**Table 3:** Case-fatality rate based on broad ICD-11 classifications.

| **ICD-11 classification** | **Number of admissions** | **Number of deaths** | **Case fatality (%)** |
| --- | --- | --- | --- |
| Certain infectious or parasitic diseases | 709,862 | 11,370 | 1.6 |
| Neoplasms | 31,464 | 1,494 | 4.8 |
| Diseases of the blood or blood-forming organs | 118,448 | 2,897 | 2.5 |
| Diseases of the immune system | 184 | 11 | 6.0 |
| Endocrine, nutritional or metabolic diseases | 86,249 | 3,709 | 4.3 |
| Mental, behavioural or neurodevelopmental disorders | 16,375 | 686 | 4.2 |
| Sleep-wake disorders | 534 | 12 | 2.25 |
| Diseases of the nervous system | 56,627 | 7,788 | 13.8 |
| Diseases of the visual system | 6,132 | 54 | 0.9 |
| Diseases of the ear or mastoid process | 7,352 | 45 | 0.6 |
| Diseases of the circulatory system | 132,590 | 6,995 | 5.3 |
| Diseases of the respiratory system | 182,472 | 6,850 | 3.8 |
| Diseases of the digestive system | 168,932 | 4,040 | 2.4 |
| Diseases of the skin | 5,527 | 117 | 2.1 |
| Diseases of the musculoskeletal system/connective tissue | 15,445 | 176 | 1.1 |
| Diseases of the genitourinary system | 139,888 | 2,505 | 1.8 |
| Pregnancy, childbirth or the puerperium | 345,977 | 804 | 0.2 |
| Certain conditions originating in the perinatal period | 146,598 | 5,659 | 3.9 |
| Developmental anomalies | 5,423 | 250 | 4.6 |
| Symptoms, signs or clinical findings, not elsewhere classified | 74,766 | 3,342 | 4.5 |
| Injury, poisoning or certain other consequences of external causes | 80,544 | 2,019 | 2.5 |
| External causes of morbidity or mortality | 25,846 | 363 | 1.4 |
| Factors influencing health status or contact with health services | 298 | 6 | 2.0 |

Similarly, hospital admissions in the Bono, Bono East, Central, Eastern, Greater Accra, Northern, Oti, Upper West, Volta and Western Regions had significantly higher odds of mortality than admissions in the Ahafo region. Admissions in the Bono region were 1.82 times more likely (AOR=1.82; 95% CI: 1.65-2.01; p<0.001), admissions in Bono East were 1.54 times more likely (AOR=1.54; 95% CI: 1.39-1.71; p<0.001), admissions in Central were 1.39 times more likely (AOR=1.39; 95% CI: 1.27-1.51; p<0.001), admissions in Eastern were 1.69 times more likely (AOR=1.69; 95% CI: 1.56-1.84; p<0.001), admissions in Greater Accra were almost two times more likely (AOR=1.90; 95% CI: 1.76-2.06; p<0.001), admissions in Northern were 1.69 times more likely (AOR=1.69; 95% CI: 1.55-1.85; p<0.001), admissions in Oti were 1.21 times more likely (AOR=1.21; 95% CI: 1.08-1.35; p=0.001), admissions in Upper West were 1.45 times more likely (AOR=1.45; 95% CI: 1.32-1.58; p<0.001), admissions in Volta were 1.74 times more likely (AOR=1.74; 95% CI: 1.60-1.89; p<0.001) and admissions in the Western region were 1.18 times more likely (AOR=1.18; 95% CI: 1.09-1.29; p<0.001) to result in mortality than admissions in the Ahafo region. However, admissions in the Ashanti region had a significantly lower odds of mortality (AOR=0.91; 95% CI: 0.84-0.99; p=0.031), and admissions in the Savannah region were 43% less likely to result in mortality (AOR=0.57; 95% CI: 0.48-0.67; p<0.001) as compared to admissions in the Ahafo region.

**Table 4:**
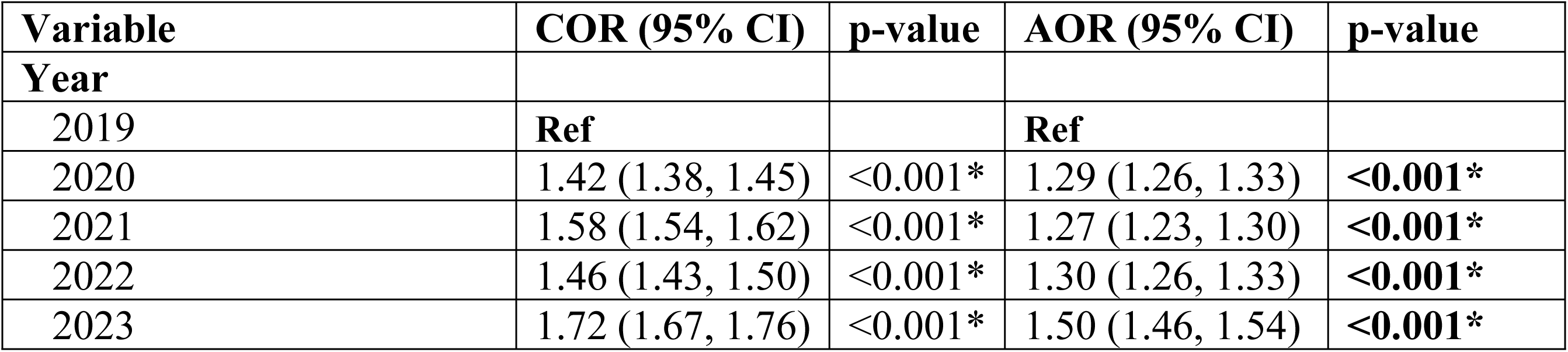
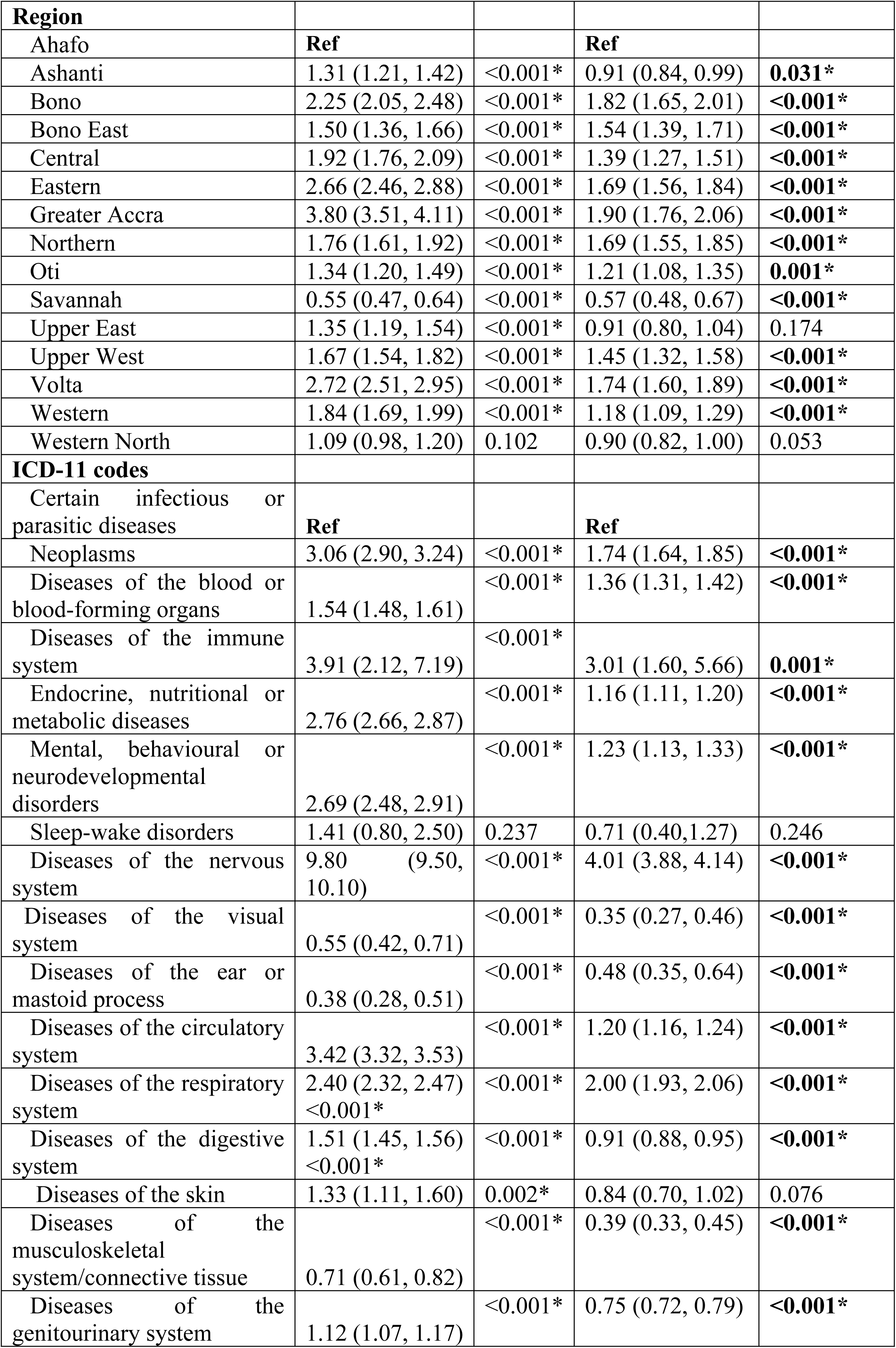
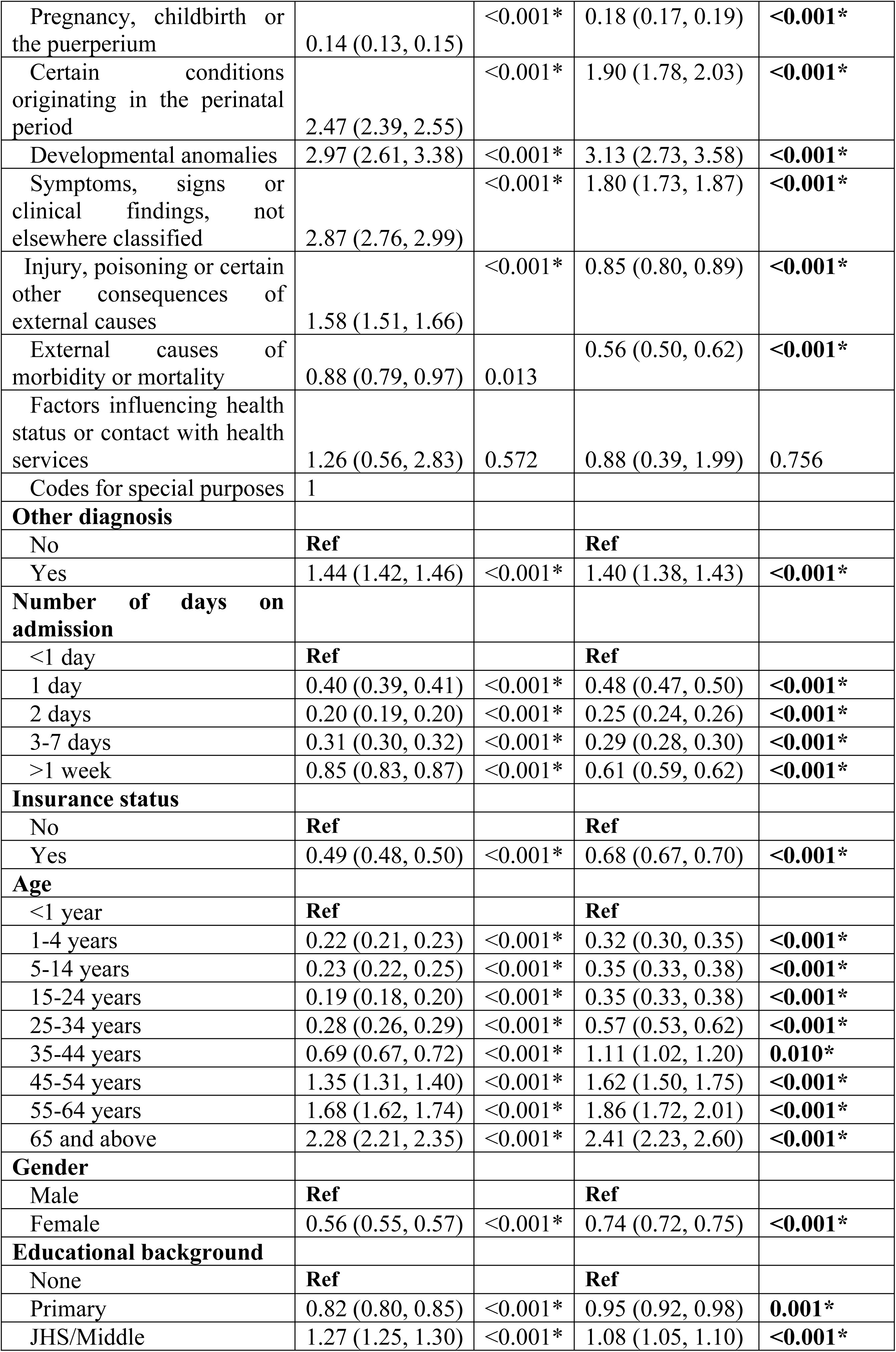
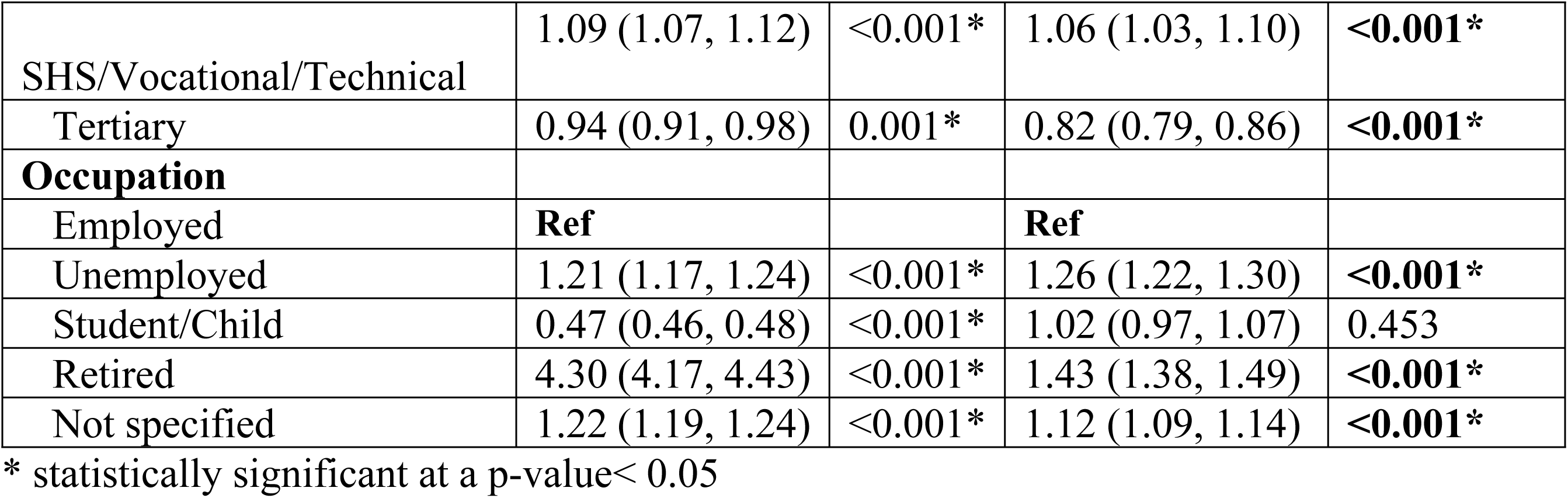
Factors associated with the odds of mortality among hospital admissions in Ghana, 2019 – 2023.

| Variable | COR (95% CI) | p-value | AOR (95% CI) | p-value |
| --- | --- | --- | --- | --- |
| **Year** |  |  |  |  |
| 2019 | **Ref** |  | **Ref** |  |
| 2020 | 1.42 (1.38, 1.45) | <0.001* | 1.29 (1.26, 1.33) | <0.001* |
| 2021 | 1.58 (1.54, 1.62) | <0.001* | 1.27 (1.23, 1.30) | <0.001* |
| 2022 | 1.46 (1.43, 1.50) | <0.001* | 1.30 (1.26, 1.33) | <0.001* |
| 2023 | 1.72 (1.67, 1.76) | <0.001* | 1.50 (1.46, 1.54) | <0.001* |

| **Region** |  |  |  |  |
| --- | --- | --- | --- | --- |
| Ahafo | **Ref** |  | **Ref** |  |
| Ashanti | 1.31 (1.21, 1.42) | <0.001* | 0.91 (0.84, 0.99) | **0.031\*** |
| Bono | 2.25 (2.05, 2.48) | <0.001* | 1.82 (1.65, 2.01) | **<0.001\*** |
| Bono East | 1.50 (1.36, 1.66) | <0.001* | 1.54 (1.39, 1.71) | **<0.001\*** |
| Central | 1.92 (1.76, 2.09) | <0.001* | 1.39 (1.27, 1.51) | **<0.001\*** |
| Eastern | 2.66 (2.46, 2.88) | <0.001* | 1.69 (1.56, 1.84) | **<0.001\*** |
| Greater Accra | 3.80 (3.51, 4.11) | <0.001* | 1.90 (1.76, 2.06) | **<0.001\*** |
| Northern | 1.76 (1.61, 1.92) | <0.001* | 1.69 (1.55, 1.85) | **<0.001\*** |
| Oti | 1.34 (1.20, 1.49) | <0.001* | 1.21 (1.08, 1.35) | **0.001\*** |
| Savannah | 0.55 (0.47, 0.64) | <0.001* | 0.57 (0.48, 0.67) | **<0.001\*** |
| Upper East | 1.35 (1.19, 1.54) | <0.001* | 0.91 (0.80, 1.04) | 0.174 |
| Upper West | 1.67 (1.54, 1.82) | <0.001* | 1.45 (1.32, 1.58) | **<0.001\*** |
| Volta | 2.72 (2.51, 2.95) | <0.001* | 1.74 (1.60, 1.89) | **<0.001\*** |
| Western | 1.84 (1.69, 1.99) | <0.001* | 1.18 (1.09, 1.29) | **<0.001\*** |
| Western North | 1.09 (0.98, 1.20) | 0.102 | 0.90 (0.82, 1.00) | 0.053 |
| **ICD-11 codes** |  |  |  |  |
| Certain infectious or parasitic diseases | **Ref** |  | **Ref** |  |
| Neoplasms | 3.06 (2.90, 3.24) | <0.001* | 1.74 (1.64, 1.85) | **<0.001\*** |
| Diseases of the blood or blood-forming organs | 1.54 (1.48, 1.61) | <0.001* | 1.36 (1.31, 1.42) | **<0.001\*** |
| Diseases of the immune system | 3.91 (2.12, 7.19) | <0.001* | 3.01 (1.60, 5.66) | **0.001\*** |
| Endocrine, nutritional or metabolic diseases | 2.76 (2.66, 2.87) | <0.001* | 1.16 (1.11, 1.20) | **<0.001\*** |
| Mental, behavioural or neurodevelopmental disorders | 2.69 (2.48, 2.91) | <0.001* | 1.23 (1.13, 1.33) | **<0.001\*** |
| Sleep-wake disorders | 1.41 (0.80, 2.50) | 0.237 | 0.71 (0.40, 1.27) | 0.246 |
| Diseases of the nervous system | 9.80 (9.50, 10.10) | <0.001* | 4.01 (3.88, 4.14) | **<0.001\*** |
| Diseases of the visual system | 0.55 (0.42, 0.71) | <0.001* | 0.35 (0.27, 0.46) | **<0.001\*** |
| Diseases of the ear or mastoid process | 0.38 (0.28, 0.51) | <0.001* | 0.48 (0.35, 0.64) | **<0.001\*** |
| Diseases of the circulatory system | 3.42 (3.32, 3.53) | <0.001* | 1.20 (1.16, 1.24) | **<0.001\*** |
| Diseases of the respiratory system | 2.40 (2.32, 2.47)<br><0.001* | <0.001* | 2.00 (1.93, 2.06) | **<0.001\*** |
| Diseases of the digestive system | 1.51 (1.45, 1.56)<br><0.001* | <0.001* | 0.91 (0.88, 0.95) | **<0.001\*** |
| Diseases of the skin | 1.33 (1.11, 1.60) | 0.002* | 0.84 (0.70, 1.02) | 0.076 |
| Diseases of the musculoskeletal system/connective tissue | 0.71 (0.61, 0.82) | <0.001* | 0.39 (0.33, 0.45) | **<0.001\*** |
| Diseases of the genitourinary system | 1.12 (1.07, 1.17) | <0.001* | 0.75 (0.72, 0.79) | **<0.001\*** |
| Pregnancy, childbirth or the puerperium | 0.14 (0.13, 0.15) | <0.001* | 0.18 (0.17, 0.19) | <0.001* |
| Certain conditions originating in the perinatal period | 2.47 (2.39, 2.55) | <0.001* | 1.90 (1.78, 2.03) | <0.001* |
| Developmental anomalies | 2.97 (2.61, 3.38) | <0.001* | 3.13 (2.73, 3.58) | <0.001* |
| Symptoms, signs or clinical findings, not elsewhere classified | 2.87 (2.76, 2.99) | <0.001* | 1.80 (1.73, 1.87) | <0.001* |
| Injury, poisoning or certain other consequences of external causes | 1.58 (1.51, 1.66) | <0.001* | 0.85 (0.80, 0.89) | <0.001* |
| External causes of morbidity or mortality | 0.88 (0.79, 0.97) | 0.013 | 0.56 (0.50, 0.62) | <0.001* |
| Factors influencing health status or contact with health services | 1.26 (0.56, 2.83) | 0.572 | 0.88 (0.39, 1.99) | 0.756 |
| Codes for special purposes | 1 |  |  |  |
| **Other diagnosis** |  |  |  |  |
| No | Ref |  | Ref |  |
| Yes | 1.44 (1.42, 1.46) | <0.001* | 1.40 (1.38, 1.43) | <0.001* |
| **Number of days on admission** |  |  |  |  |
| <1 day | Ref |  | Ref |  |
| 1 day | 0.40 (0.39, 0.41) | <0.001* | 0.48 (0.47, 0.50) | <0.001* |
| 2 days | 0.20 (0.19, 0.20) | <0.001* | 0.25 (0.24, 0.26) | <0.001* |
| 3-7 days | 0.31 (0.30, 0.32) | <0.001* | 0.29 (0.28, 0.30) | <0.001* |
| >1 week | 0.85 (0.83, 0.87) | <0.001* | 0.61 (0.59, 0.62) | <0.001* |
| **Insurance status** |  |  |  |  |
| No | Ref |  | Ref |  |
| Yes | 0.49 (0.48, 0.50) | <0.001* | 0.68 (0.67, 0.70) | <0.001* |
| **Age** |  |  |  |  |
| <1 year | Ref |  | Ref |  |
| 1-4 years | 0.22 (0.21, 0.23) | <0.001* | 0.32 (0.30, 0.35) | <0.001* |
| 5-14 years | 0.23 (0.22, 0.25) | <0.001* | 0.35 (0.33, 0.38) | <0.001* |
| 15-24 years | 0.19 (0.18, 0.20) | <0.001* | 0.35 (0.33, 0.38) | <0.001* |
| 25-34 years | 0.28 (0.26, 0.29) | <0.001* | 0.57 (0.53, 0.62) | <0.001* |
| 35-44 years | 0.69 (0.67, 0.72) | <0.001* | 1.11 (1.02, 1.20) | 0.010* |
| 45-54 years | 1.35 (1.31, 1.40) | <0.001* | 1.62 (1.50, 1.75) | <0.001* |
| 55-64 years | 1.68 (1.62, 1.74) | <0.001* | 1.86 (1.72, 2.01) | <0.001* |
| 65 and above | 2.28 (2.21, 2.35) | <0.001* | 2.41 (2.23, 2.60) | <0.001* |
| **Gender** |  |  |  |  |
| Male | Ref |  | Ref |  |
| Female | 0.56 (0.55, 0.57) | <0.001* | 0.74 (0.72, 0.75) | <0.001* |
| **Educational background** |  |  |  |  |
| None | Ref |  | Ref |  |
| Primary | 0.82 (0.80, 0.85) | <0.001* | 0.95 (0.92, 0.98) | 0.001* |
| JHS/Middle | 1.27 (1.25, 1.30) | <0.001* | 1.08 (1.05, 1.10) | <0.001* |
| SHS/Vocational/Technical | 1.09 (1.07, 1.12) | <0.001* | 1.06 (1.03, 1.10) | <0.001* |
| Tertiary | 0.94 (0.91, 0.98) | 0.001* | 0.82 (0.79, 0.86) | <0.001* |
| **Occupation** |  |  |  |  |
| Employed | **Ref** |  | **Ref** |  |
| Unemployed | 1.21 (1.17, 1.24) | <0.001* | 1.26 (1.22, 1.30) | <0.001* |
| Student/Child | 0.47 (0.46, 0.48) | <0.001* | 1.02 (0.97, 1.07) | 0.453 |
| Retired | 4.30 (4.17, 4.43) | <0.001* | 1.43 (1.38, 1.49) | <0.001* |
| Not specified | 1.22 (1.19, 1.24) | <0.001* | 1.12 (1.09, 1.14) | <0.001* |

\* statistically significant at a p-value< 0.05

Moreover, hospital admissions due to diseases of the digestive system, diseases of the musculoskeletal system/connective tissue, diseases of the genitourinary system, pregnancy, childbirth or the puerperium, diseases of the visual system, diseases of the ear or mastoid process, injury, poisoning or inevitable other consequences of external causes and external causes of morbidity or mortality had significantly lower odds of mortality than certain infectious or parasitic diseases. However, hospital admissions due to neoplasms, diseases of the blood or blood-forming organs, diseases of the immune system, endocrine, nutritional or metabolic diseases, mental, behavioural or neurodevelopmental disorders, diseases of the nervous system, diseases of the circulatory system, diseases of the respiratory system, certain conditions originating in the perinatal period, developmental anomalies and symptoms, signs or clinical findings, not elsewhere classified had significantly higher odds of mortality as compared to certain infectious or parasitic diseases.

Hospital admissions from diseases of the digestive system were 9% less likely (AOR=0.91; 95% CI: 0.88-0.95; p<0.001), diseases of the musculoskeletal system/connective tissue were 61% less likely (AOR=0.39; 95% CI: 0.33-0.45; p<0.001), diseases of the genitourinary system were 25% less likely (AOR=0.75; 95% CI: 0.72-0.79; p<0.001), pregnancy, childbirth or the puerperium conditions were 82% less likely (AOR=0.18; 95% CI: 0.17-0.19; p<0.001), diseases of the visual system were 65% less likely (AOR=0.35; 95% CI: 0.27-0.46;p<0.001), diseases of the ear or mastoid process were 52% less likely (AOR=0.48; 95% CI: 0.35-0.64; p<0.001), injury, poisoning or inevitable other consequences of external causes were 15% less likely (AOR=0.85; 95% CI: 0.80-0.89; p<0.001) and external causes of morbidity or mortality were 44% less likely (AOR=0.56; 95% CI: 0.50-0.62; p<0.001) to result in mortality as compared to certain infectious or parasitic diseases.

Additionally, hospital admissions due to neoplasms were 1.74 times more likely (AOR=1.74; 95% CI: 1.64-1.85; p<0.001), diseases of the blood or blood-forming organs were 1.36 more likely (AOR=1.36; 95% CI: 1.31-1.42; p<0.001), diseases of the immune system were three times more likely (AOR=3.01; 95% CI: 1.60-5.66; p=0.001), endocrine, nutritional or metabolic diseases were 1.16 times more likely (AOR=1.16, 95% CI: 1.11-1.20; p<0.001), mental, behavioural or neurodevelopmental disorders were 1.23 times more likely (AOR=1.23; 95% CI: 1.13-1.33; p<0.001), diseases of the nervous system were four times more likely (AOR=4.01; 95% CI: 3.88-4.14; p<0.001), diseases of the circulatory system were 1.2 times more likely (AOR=1.20; 95% CI: 1.16-1.24; p<0.001), diseases of the respiratory system were twice more likely (AOR=2.00; 95% CI: 1.93-2.06; p<0.001), certain conditions originating in the perinatal period were almost two times more likely (AOR=1.90; 95% CI: 1.78-2.03; p<0.001), developmental anomalies were three times more likely (AOR=3.13; CI: 2.73-3.58; p<0.001) and symptoms, signs or clinical findings, not elsewhere classified were 1.8 times more likely (AOR=1.80; 95% CI: 1.73-1.87; p<0.001) to lead to mortality as compared to certain infectious or parasitic diseases.

Furthermore, hospital admissions with another diagnosis apart from the primary diagnosis had higher odds of mortality than those without another diagnosis (AOR=1.40; 95% CI: 1.38-1.43; p<0.001). Furthermore, persons who spent one day on admission were 52% less likely result in mortalities (AOR=0.48; 95% CI: 0.47-0.50; p<0.001), persons who spent two days on admission were 75% less likely (AOR=0.25; 95% CI: 0.24-0.26; p <0.001), persons who spent 3 to 7 days on admission were 71% less likely (AOR=0.29; 95% CI: 0.28-0.30; p<0.001) and persons who spent more than one week on admission were 39% less likely (AOR=0.61; 95% CI: 0.59-0.62; p<0.001) to become mortalities as compared to persons who spent less than one day on admission.

Additionally, individuals with insurance were 32% less likely to experience mortality compared to those without insurance (AOR = 0.68, CI = 0.67-0.70, p < 0.001). Also, persons aged 1-34 years had significantly lower odds of mortality than persons aged less than one year. However, persons aged 35 years and above had considerably higher odds of mortality than persons aged less than one year. Persons aged 1-4 years were 68% less likely (AOR=0.32; 95% CI: 0.30-0.35; p<0.001), persons aged 5-14 years were 65% less likely (AOR=0.35; 95% CI: 0.33-0.38; p<0.001), persons aged 15-24 years were also 65% less likely (AOR=0.35; 95% CI: 0.33-0.38; p<0.001) and persons aged 25-34 years were 43% less likely (AOR=0.57; 95% CI: 0.53-0.62; p<0.001) to result in mortality as compared to persons aged less than one year. On the contrary, persons aged 35-44 years were 1.11 times more likely (AOR= 1.11; 95% CI: 1.02-1.20; p=0.010), persons aged 45-54 years were 1.62 times more likely (AOR=1.62; 95% CI: 1.50-1.75; p<0.001), persons aged 55-64 years were almost twice more likely (AOR=1.86; 95% CI: 1.72-2.01; p<0.001) and persons aged 65 years and above were more than twice more likely (AOR=2.41; 95% CI: 2.23-2.60; p<0.001) to result in mortality as compared to persons aged less than one year.

Similarly, females admitted to the hospital had lower odds of mortality compared to males (AOR = 0.74; 95% CI: 0.72-0.75; p < 0.001). Regarding educational background, individuals with primary and tertiary education had lower odds of mortality than those with no education. Moreover, JHS/Middle school and SHS/Vocational/Technical had higher odds of mortality. Persons with primary education were 5% less likely (AOR=0.95; 95% CI: 0.92-0.98; p-value=0.001), and persons with tertiary education were 18% less likely (AOR=0.82; 95% CI: 0.79-0.86; p<0.001) to result in mortality as compared to persons with no education. Persons with JHS/Middle school education were however, 1.08 times more likely (AOR= 1.08: 95% CI: 1.05, 1.10; p<0.001 and persons with SHS/Vocational/Technical education were 1.06 times more likely (AOR=1.06; 95% CI: 1.03-1.10; p<0.001) to result in mortality as compared to persons with no education.

Finally, unemployed persons (AOR=1.26; 95% CI: 1.22, 1.30; p<0.001), those who were retired (AOR=1.43; 95% CI: 1.38-1.49; p<0.001), and those whose occupation was not specified (AOR=1.12: 95% CI: 1.09-1.14; p<0.001) had significantly higher odds of mortality than those who were employed.

## Discussion

This study examined Ghana’s national hospital admissions and death patterns from 2019 to 2023 using a reliable dataset of over 2.3 million hospital admissions. The study offers significant implications for public health policy and healthcare delivery by highlighting crucial demographic, regional, and clinical characteristics linked to hospital mortality through the use of ICD-11 classifications and multivariate logistic regression analysis.

### Trends in Morbidity and Mortality

Over the study period, the general distribution of hospital admissions in Ghana exhibited a declining trend, peaking in 2019 and reaching its lowest point in 2023. It is interesting to note that the odds of mortality rose year-on-year, with admissions in 2023 being 1.50 times more likely to result in death than those in 2019. These patterns could be partially explained by the COVID-19 pandemic’s impact on healthcare services, which has been linked to higher mortality rates as a result of overburdened health systems, delayed care access, and delayed treatment of both acute and chronic illnesses [16], [17], [18], [19], [20].

### Regional disparities in mortality

Notable geographical differences were found, with the highest mortality odds observed in the Greater Accra, Eastern, and Volta regions. In Ghana and other sub-Saharan African nations, these results are in line with research that shows regional disparities in healthcare access and resource distribution [21], [22], [23]. Despite often having more resources, urban locations may still have higher mortality rates due to larger patient volumes and the referral of complex patients from peripheral facilities. On the other hand, regions like Savannah and Ashanti recorded lower odds, which might be due to either improved care delivery or fewer people reporting urgent illnesses.

### Disease burden and case fatality rates

According to the ICD-11 categorisation, the study found that parasitic and infectious disorders accounted for 30.1% of hospital admissions, with a case-fatality rate of only 1.6%. This implies that, with timely care, many illnesses are usually treatable. In contrast, diseases of the nervous system, circulatory system, and respiratory system, although less frequent, were associated with significantly higher case fatality rates (13.8%, 5.3%, and 3.8%, respectively). These results are consistent with international research that highlights the mortality linked to neurological and cardiovascular disorders because of their rapid advancement and the complexity of care needed [24], [25]. It also reaffirms literature emphasising the critical nature of neurological conditions, which often present late and require specialised care that may not be widely available in many healthcare facilities [26], [27]. Other causes with high case fatality included diseases of the immune system (6.0%), circulatory system (5.3%), and neoplasms (4.8%). The significantly lower mortality associated with admission for pregnancy and childbirth (0.2%) may reflect improvements in maternal healthcare services and interventions under Ghana’s reproductive health programs. However, persistent disparities across other disease categories highlight the need for targeted health system strengthening [28], [29].

Additionally, certain conditions originating in the perinatal period and developmental anomalies showed higher odds of mortality, showing vulnerabilities among neonates and children with congenital conditions. These results support the need for improved maternity and newborn care services, which, when properly funded, are successful in lowering early-life mortality [30], [31], [32].

### Sociodemographic factors influencing mortality

Age, sex, education, and employment status were significant determinants of mortality. Older adults (65+) had the highest odds of mortality, which aligns with broader epidemiological evidence showing age as a primary risk factor for poor outcomes in hospitalised patients [33], [34]. Males were more likely to die than females, which could be attributed to differences in health-seeking behaviour, biological factors, and comorbidity patterns [35], [36], [37].

Mortality and educational attainment had a complex relationship. Middle and secondary education levels were associated with higher mortality rates compared to no formal education, whereas postsecondary education was protective. Although this conclusion merits more qualitative investigation, it may be the result of occupational risks or lack of consistent health information among these populations [38], [39].

Another essential element was employment status: those who were unemployed or retired had greater death probabilities. This bolsters the body of research that shows socioeconomic inequality raises health risks because it restricts access to healthcare and makes people more susceptible to stress-related illnesses [40].

### Health System and Insurance Coverage

Notably, patients with health insurance had a significantly lower mortality rate than those without, highlighting the contribution of Ghana’s National Health Insurance Scheme (NHIS) to improved access to prompt medical care. This is consistent with earlier studies that found insurance coverage is associated with better health outcomes and higher healthcare consumption in LMICs [41], [42], [43], [44].

These findings support the need for public education on early healthcare seeking and the development of effective triage systems. The length of hospitalisation also showed a strong association with mortality, with longer stays being protective, possibly due to sustained care and clinical stabilisation. In comparison, shorter stays were associated with a higher risk of death, indicating that many fatalities occurred among critically ill patients who presented late to the hospital [45], [46].

### Clinical complexity and comorbidity

Patients with additional diagnoses had higher odds of mortality, highlighting the challenge of managing multidisciplinary care, which may not be readily available in all hospitals [47]. This emphasises the need for integrated care pathways and improved diagnostic capacities at all levels of care.

### Implications for Policy and Practice

Investments in critical care services and diagnostic capabilities are desperately needed, as evidenced by the high fatality rates linked to specific ICD-11 categories, including neurological, circulatory, and respiratory illnesses. Reexamining training and resource distribution is also necessary, particularly in underprivileged areas where mortality rates are disproportionate.

The results also confirm the value of health insurance and education in lowering the chance of death. Therefore, increasing NHIS coverage and integrating public health education within the health system should be top priorities for policymakers.

## Conclusion

This study offers an in-depth examination of Ghana’s hospital mortality trends and their contributing factors. Despite progress in healthcare access and service delivery, the national case fatality rate of 2.6% highlights persistent gaps, especially for conditions with high clinical complexity, such as neurological, circulatory, and respiratory diseases.

Improving hospital outcomes nationwide will require investments in community-based health education, rapid diagnosis, and critical care capacity. To reduce regional inequities, strengthen the NHIS, and enhance treatment for high-risk groups, including the elderly and patients with complex medical needs, the findings emphasise the necessity of focused policy initiatives. Strengthening health system responsiveness, particularly in high-burden regions and among vulnerable populations, is essential to improving health outcomes and reducing avoidable hospital deaths.

### Recommendations

To lower hospital-based mortality in Ghana, the following policy proposals are put forth in light of the study’s findings:

### Enhance critical care capacity

The Ghana Health Service (GHS) and the Ministry of Health (MoH) should invest in modernising critical care facilities, particularly in areas with high mortality rates. This involves providing ventilators, specialised personnel, and intensive care units (ICUs) to hospitals.

### Increase and broaden NHIS coverage

To alleviate financial burdens on care, the National Health Insurance Authority (NHIA) should intensify efforts to enrol and renew members, particularly focusing on vulnerable groups and workers in the informal sector.

### Improve regional health equity

Disparities in healthcare supply chains, health personnel, and infrastructure must be addressed by regional health directorates (government). Investment in underperforming areas, such as the Eastern and Volta regions, must be prioritised.

### Enhance data quality and surveillance

To ensure accurate ICD coding and comprehensive reporting, including from underrepresented regions such as the Northeast, the GHS’s Health Information Management Systems (HIMS) unit should strengthen hospital data systems.

### Encourage early health-seeking behaviour

The GHS’s Health Promotion Division ought to spearhead national initiatives that highlight the importance of early health-seeking behaviour, particularly for acute and chronic illnesses.

### Implement integrated multimorbidity care models

With assistance from the Ghana College of Physicians and Surgeons, teaching hospitals, regional, and district health facilities should be trained to adopt and implement multidisciplinary care techniques for patients with comorbidities.

### Targeted geriatric and perinatal interventions

To address the high mortality burden in these age groups, the MoH should establish national protocols centred on geriatric and newborn care, allocating resources and providing specialised training.

### Limitations

This study has certain limitations, despite the sizable and nationally representative dataset. Initially, the analysis was limited to data from hospitals and may have missed deaths that occurred outside of medical institutions, potentially resulting in an underestimation of mortality. Second, coding errors and misclassification bias can arise when ICD-11 classifications and administrative data are relied upon, particularly in facilities with limited resources. Third, the capacity to evaluate case severity and clinical pathways leading to mortality is limited by the absence of detailed clinical data (e.g., laboratory results, comorbidities beyond the second diagnosis). Furthermore, socioeconomic variables that might have had a significant impact on the results but were excluded included housing, income, and living in an urban or rural area. Lastly, the statistics for the North East Region were not included because they were insufficient, which would have impacted the broad applicability of the results.

## Declarations

### Ethics approval and consent to participate

To extract and use the pertinent datasets, the Ghana Health Service’s Policy Planning, Monitoring and Evaluation Division was consulted for the necessary authorisation. The dataset used for analysis does not include any identifying information about the study participants. The Ghana Health Service Ethics Review Committee granted ethical approval before the commencement of the study, and confidentiality was maintained throughout.

### Consent for publication

Not applicable

### Availability of data and materials

The datasets generated and analysed during this study are not fully available but can be made available upon reasonable request to the corresponding author.

